# An Informatics Consult approach for generating clinical evidence for treatment decisions

**DOI:** 10.1101/2021.01.10.21249331

**Authors:** Alvina G. Lai, Wai Hoong Chang, Constantinos A. Parisinos, Michail Katsoulis, Ruth M. Blackburn, Anoop D. Shah, Vincent Nguyen, Spiros Denaxas, George Davey Smith, Tom R. Gaunt, Krishnarajah Nirantharakumar, Murray P. Cox, Donall Forde, Folkert Asselbergs, Steve Harris, Sylvia Richardson, Reecha Sofat, Richard J.B. Dobson, Aroon Hingorani, Riyaz Patel, Jonathan Sterne, Amitava Banerjee, Alastair K. Denniston, Simon Ball, Neil J. Sebire, Nigam H. Shah, Graham R. Foster, Bryan Williams, Harry Hemingway

**Affiliations:** Institute of Health Informatics, University College London, London, UK; Health Data Research UK, UK; University College London Hospitals NIHR Biomedical Research Centre, London, UK; The Alan Turing Institute, London, UK; Medical Research Council Integrative Epidemiology Unit, University of Bristol, Bristol, UK; Population Health Sciences. Bristol Medical School, University of Bristol, Bristol, UK; Institute of Applies Health Research, University of Birmingham, UK; Statistics and Bioinformatics Group, School of Fundamental Sciences, Massey University, New Zealand; Public Health Wales, University Hospital of Wales, Cardiff, UK; Department of Cardiology, University Medical Center Utrecht, Utrecht University, Utrecht, Netherlands; Institute of Cardiovascular Science, University College London, London, UK; University College London Hospitals NHS Trust, London, UK; Medical Research Council Biostatistics Unit, University of Cambridge, Cambridge, UK; Department of Biostatistics and Health Informatics, King’s College London, London, UK; Barts Health NHS Trust, The Royal London Hospital, Whitechapel Rd, London, UK; University Hospitals Birmingham NHSFT, Birmingham, UK; UCL Great Ormond Street Institute of Child Health, London, UK; NIHR Great Ormond Street Hospital Biomedical Research Centre, London, UK; Department of Medicine, School of Medicine, Stanford University, Stanford California, USA; Barts Liver Centre, Blizard Institute, Queen Mary University of London, London, UK

## Abstract

An Informatics Consult has been proposed in which clinicians request novel evidence from large scale health data resources, tailored to the treatment of a specific patient, with return of results in clinical timescales. However, the availability of such consultations is lacking. We seek to provide an Informatics Consult for a situation where a treatment indication and contraindication coexist in the same patient, i.e., anti-coagulation use for stroke prevention in a patient with both atrial fibrillation (AF) and liver cirrhosis. We examined four sources of evidence for the effect of warfarin on stroke risk (efficacy) or all-cause mortality (safety) from: (i) randomised controlled trials (RCTs), (ii) meta-analysis of prior observational studies, (iii) trial emulation (using population electronic health records (N = 3,854,710) and (iv) genetic evidence (Mendelian randomisation). We developed prototype forms to request an Informatics Consult and return of results in electronic health record systems. We found 0 RCT reports and 0 trials recruiting for patients with AF and cirrhosis. We found broad concordance across the three new sources of evidence we generated. Meta-analysis of prior observational studies showed that warfarin use was associated with lower stroke risk (hazard ratio [HR] = 0.71). In a target trial emulation, warfarin was associated with lower all-cause mortality (HR = 0.61) and ischaemic stroke (HR = 0.27). Mendelian randomisation served as a drug target validation where we found that lower levels of vitamin K1 (warfarin is a vitamin K1 antagonist) are associated with lower stroke risk. A pilot survey with an independent sample of 34 clinicians revealed that 85% of clinicians found information on prognosis useful and that 79% thought that they should have access to the Informatics Consult as a service within their healthcare systems. We identified candidate steps for automation to scale evidence generation and to accelerate the return of results within clinical timescales.

## Introduction

Evidence informing treatment decisions traditionally takes years to generate and leaves many clinical uncertainties unaddressed[1]. Especially in patients with two or more conditions (multimorbidity), it has been hard to generate evidence tailored to ‘patients like me’ and embed this evidence in clinical decision making using electronic health records. There are thousands of clinical practice recommendations, but only a small proportion (15-20%) of these recommendations are supported by level A (trial) evidence[1–6]. A systematic review of trial registration records found that 79% of randomised controlled trials (RCTs) excluded patients with concomitant chronic conditions[6]. The United States Food and Drug Administration, Medicines Healthcare Regulatory Authority and European Medicines Agency are increasingly recognising the role of real-world evidence[7] but guidance thus far has not considered its near real-time generation.

The Informatics Consult concept has been proposed[8–11] to produce on-demand evidence in which clinicians request novel evidence based on the care of prior patients to inform the treatment of a specific patient, with return of results in decision-relevant clinical timescales. For example, a hepatologist seeing a patient with cirrhosis in the clinic learns that they have developed atrial fibrillation. What evidence is, or could rapidly be, available to inform a decision on anti-coagulation for stroke prevention? This is an example of a treatment indication and a treatment contra-indication coexisting in the same patient. Initial experience of the Informatics Consult from Stanford University, has focussed on questions of prognosis (rather than treatment decisions) and highlighted analytical and scaling challenges in returning results in clinical timescales[11]. However, demonstrators of the Informatics Consult for treatment decisions are lacking.

Our objective was to demonstrate proof of concept of the Informatics Consult in clinically actionable timescales using the atrial fibrillation-cirrhosis-warfarin example. Specifically, we sought to (i) develop prototype electronic health record forms for requesting an Informatics Consult and return of results, (ii) generate four sources of evidence for an Informatics Consult (evaluate available RCT evidence, meta-analyse prior observational studies, emulate a target trial using electronic health records, and Mendelian randomisation[12,13]), (iii) for each form of evidence, to identify steps necessary for automation to accelerate evidence generation and return of results to clinicians, and scale across multiple exemplars (Table S1) and (iv) explore clinician acceptability of the Informatics Consult.

## Methods

### Developing prototype electronic health record (EHR) request form and report form

An Informatics Consult is triggered by a request made by a clinician from the electronic health records (EHR) for multiple novel sources of evidence. We developed prototypes for the request form and the report form in consultation with the Chief Clinical Research Informatics Officer and clinicians (cardiologists, hepatologists and clinical pharmacologists). We illustrate possible forms based on an EHR platform, but similar design principles are relevant in EHRs from other vendors, and in other settings, including primary care.

### Retrieving information on currently recruiting trials

We searched for previously reported and currently recruiting trials on anticoagulants in patients with atrial fibrillation, with stroke or mortality in the primary outcome using the ClinicalTrials.gov registry. We then used the 8-digit National Clinical Trial numbers to retrieve detailed information on inclusion and exclusion criteria for each trial to identify if any trials included patients with both atrial fibrillation and cirrhosis.

### Meta-analysis of prior observational studies

*<u>Study identification:</u>* We searched PubMed for peer-reviewed articles using the keywords “antithrombotic”, “anticoagulant”, “warfarin”, “cirrhosis” and “atrial fibrillation”. We considered eligible studies as those reporting the effects of anticoagulation therapy in patients with both liver cirrhosis and atrial fibrillation. We excluded reviews, single case reports, editorials and small case series (< 10 cases). *<u>Data extraction:</u>* We extracted the following variables: author, setting, eligibility criteria, number of patients with atrial fibrillation and cirrhosis, number of patients in treated and untreated groups and summary measures. Analyses were performed following PRISMA guidelines. Outcomes of interest were mortality and ischaemic stroke. *<u>Statistical analysis</u>*. A meta-analysis of associations was performed by pooling hazard ratios (HRs) or odds ratios (ORs) depending on data availability from observational studies using DerSimonian and Laird random-effects models. We also performed leave-one-out sensitivity analyses.

### Target trial emulation

We used population-based EHRs to perform a target trial emulation, which is the application of design principles from RCTs to inform analyses on observational data[14,15]. We obtained informational governance approval from the Medicines Healthcare Regulatory Authority (UK) Independent Scientific Advisory Committee (20_078R) to analyse the Clinical Practice Research Datalink (CPRD) linked to secondary care Hospital Episode Statistics and the Office for National Statistic death registration. The study population was 3,854,710 adults aged ≥ 30 years. Phenotype definitions for atrial fibrillation, cirrhosis and other conditions included as baseline covariates as well as definitions for prescriptions are available at https://caliberresearch.org/portal and have previously been validated[16,17]. Phenotypes for primary care records were generated using Read clinical terminology (version 2). Phenotypes for secondary care records were generated using ICD-10 terms.

We developed a target trial protocol where eligibility criteria, treatment assignment, treatment strategy, follow-up period, causal contrast and statistical analyses were specified. Each component of the trial protocol is matched as closely as possible to the design of a randomised trial with minor modifications to accommodate the use of observational data. We employed the intention-to-treat effect as a causal contrast, which was warfarin initiation versus no initiation at baseline. To emulate a target trial, we ensured that individuals are classified as warfarin initiators versus non-initiators at baseline (i.e., using baseline information to assign baseline treatment status). To perform the intention-to-treat analysis, we assigned individuals to the initiator group if they use warfarin within 3 months of the baseline date. The baseline is defined as the latest date by which a patient has both cirrhosis and atrial fibrillation given that all eligibility criteria are met. As we were interested in assessing the effects of warfarin use on stroke, we have also excluded prevalent cases of ischaemic stroke. Individuals were followed until the development of an outcome of interest, which were all- cause mortality and incident ischaemic stroke. Propensity score matching (PSM) analyses were performed to reduce bias by matching the warfarin initiator and non-initiator groups. PSM was performed using the nearest-neighbour matching method (a 1:3 match was performed where possible) with a calliper width of 0.2 of the standard deviation of the logit of the propensity score. The PSM cohort was subjected to analyses of all-cause mortality and incident stroke using the Kaplan-Meier and logrank test method and the Cox proportional hazard regression model. As patients and clinicians would be interested in understanding risk in specific demographic categories, we performed subgroup analyses for all-cause mortality in patients aged ≤ 65, aged > 65, men, women and in patients with normal international normalise ratio (INR) measurements.

### Genetic evidence: Two-sample Mendelian Randomisation (MR)

As an example of drug target validation in the general population, we performed MR to investigate the causal relationship between warfarin use and stroke risk. Vitamin K1 (phylloquinone) is a central component in the production of blood coagulation factors. Warfarin (a vitamin K antagonist) inhibits the activity of vitamin K epoxide reductase to interfere with the recycling of vitamin K and to reduce blood clotting. We considered four single nucleotide polymorphisms (SNPs) that predict circulating phylloquinone (vitamin K1) selected from a genome-wide meta-analysis study on Europeans[18]. Four SNPs, all on separate chromosomes, were selected as they had the strongest association with circulating phylloquinone: rs2108622 (chromosome 19), rs2192574 (chromosome 2), rs4645543 (chromosome 8) and rs6862071 (chromosome 5). We retrieved genome wide association study (GWAS) summarised data for stroke outcomes from the MEGASTROKE study[19]. MR was performed using the “MendelianRandomisation” package in R[20]. We explored four methods for MR; inverse-variance weighted (IVW), MR-Egger, simple median and weighted median.

### Potential for automating the Informatics Consult

We tasked a review panel with expertise in health informatics, epidemiology, evidence synthesis and computer science (drawn from co-authors) with two questions: 1) What are the most time-consuming tasks for each of the 4 streams of evidence identification and generation? (2) What automation opportunities might be important for acceleration and scaling.

### Clinician acceptability of the Informatics Consult

After the initial development of the Consult, to gain insights into the acceptability and feasibility of the Informatics Consult, we conducted a pilot survey with an independent sample of 34 clinicians who had not taken part in the research.

## Results

### Prototype EHR request form

Clinicians wanted the request form (Figure 1) to include a succinct free text statement of the clinical question, auto propagation of the diagnosis combination from the patient’s EHR, and suggested structured treatment, efficacy and safety outcomes, based on inputs from the survey. Some clinicians wanted to further specify eligibility criteria for target trial emulation. Interestingly clinicians wanted not only evidence directly related to treatment effectiveness, but also requested national prevalence estimates, prognosis, and current treatment variation.

**Figure 1.**
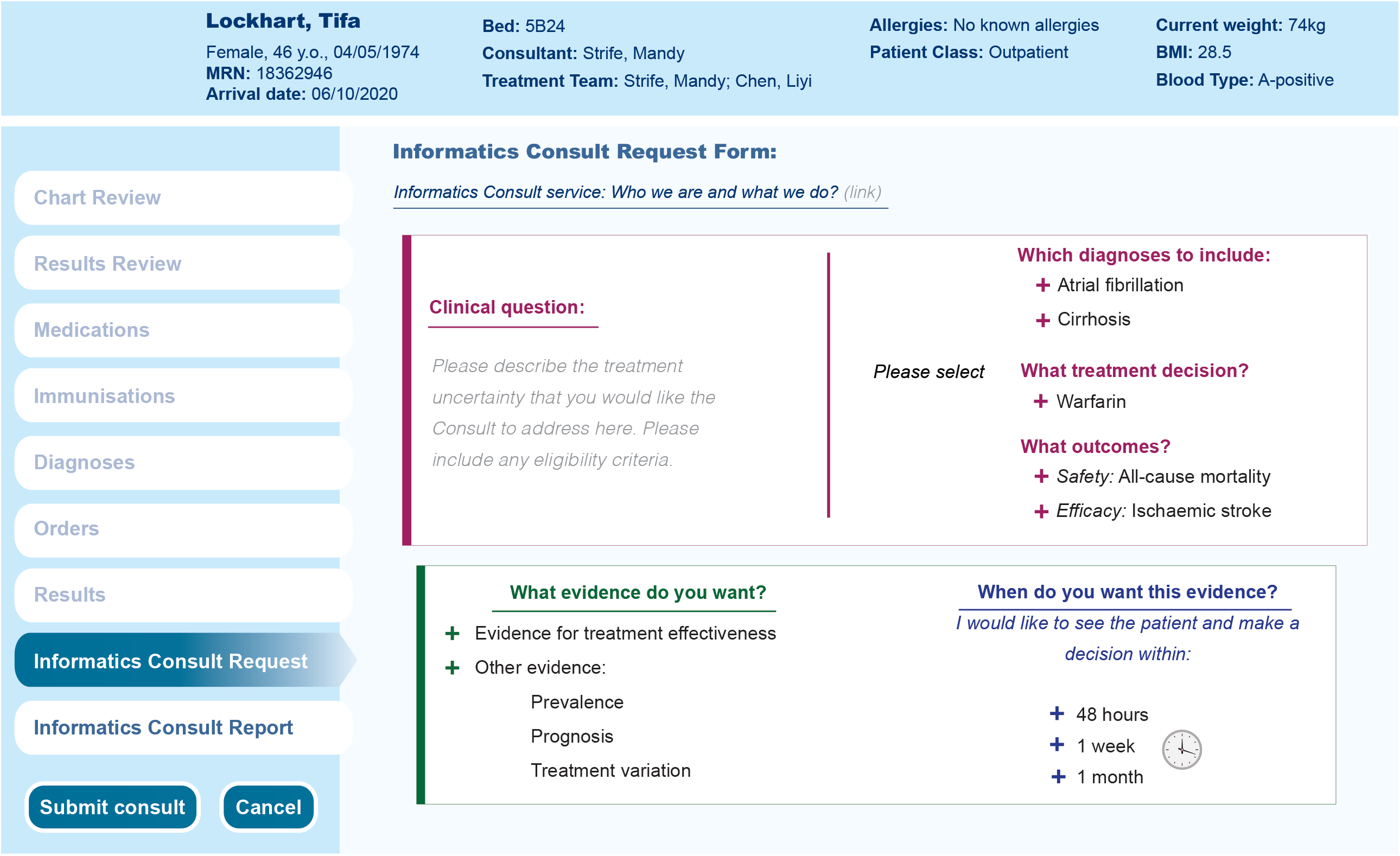
Electronic health record Informatics Consult request form prototype

### Prototype EHR report

We provide an overall summary report based on new evidence on warfarin use (lower all-cause mortality and lower stroke risk) in patients with atrial fibrillation and cirrhosis (Figure 2). Through the Consult report, clinicians will have the opportunity to queue patients for RCTs where relevant. We summarise evidence on prevalence, 1-year background mortality risk, meta-analysis of observational studies, target trial emulation results and genetic evidence. The detailed results included in the report for each form of evidence were provided below.

**Figure 2.**
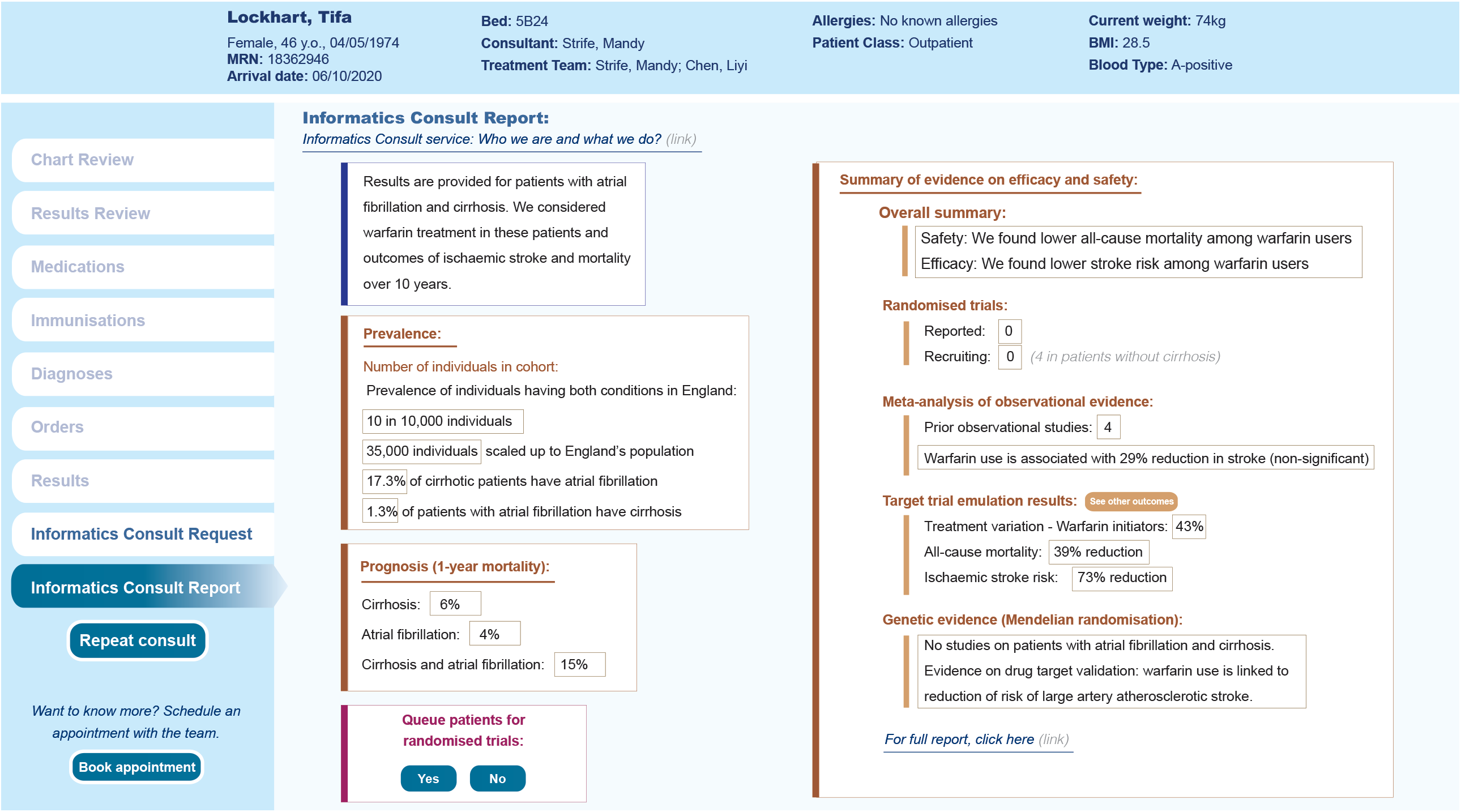
Electronic health record Informatics Consult report prototype.

### Reported and currently recruiting randomised trials

We did not identify any previous reported RCTs on anticoagulants relevant to patients with cirrhosis. We identified four currently recruiting anticoagulant trials. All four trials reported exclusion criteria related to liver cirrhosis such as contraindication to anticoagulation, i.e., hepatic impairment and elevated liver function tests (Figure 3).

**Figure 3.**
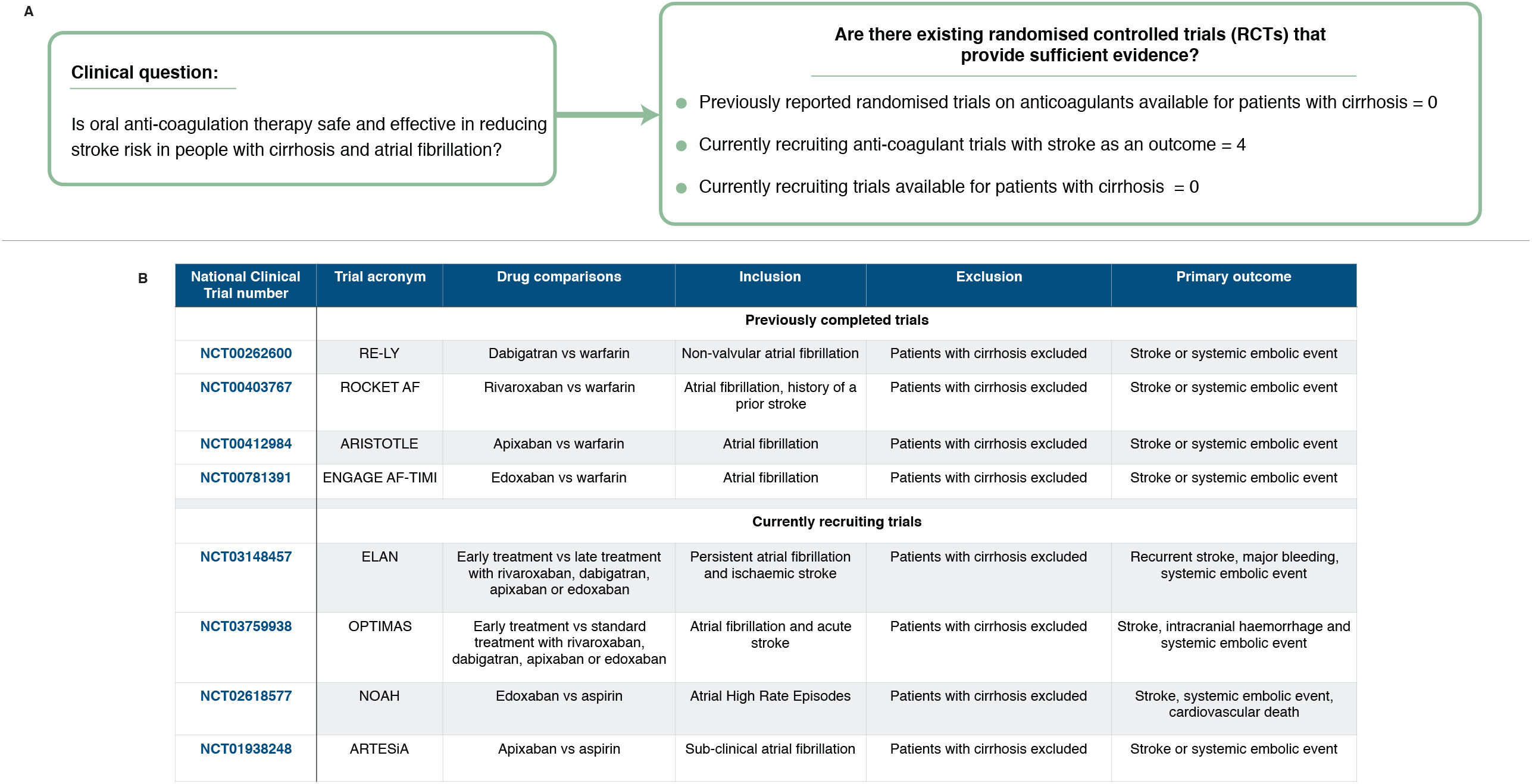
Informatics Consult Report: Detailed trial evidence and currently recruiting trials of anticoagulation in patients with atrial fibrillation and cirrhosis to reduce stroke risk. (A) Clinical question and summary of trial evidence. (B) Previously completed and currently recruiting randomised trials evaluating anticoagulants and stroke outcomes have exclusion criteria related to cirrhosis.

### Meta-analysis of observational studies on warfarin use and ischaemic stroke

We identified 142 articles from PubMed, of which only 4 observational studies remained eligible on full-text review, and were included in the meta-analysis[21–24] (Figure 4A). The pooled hazard ratio (HR) of warfarin use in patients with atrial fibrillation and cirrhosis on stroke was 0.71, 95% confidence interval [CI] (CI = 0.39 – 1.29), with high heterogeneity between studies I^2^ = 73% (Figure 4B). We also performed the leave-one-out sensitivity analysis with HRs ranging from 0.54 (CI = 0.30 - 1.00) to 0.92 (CI = 0.55 - 1.54). Only 1 study reported warfarin use and all-cause mortality and found a lower mortality risk (HR = 0.65; = 0.55 – 0.76)[21].

**Figure 4.**
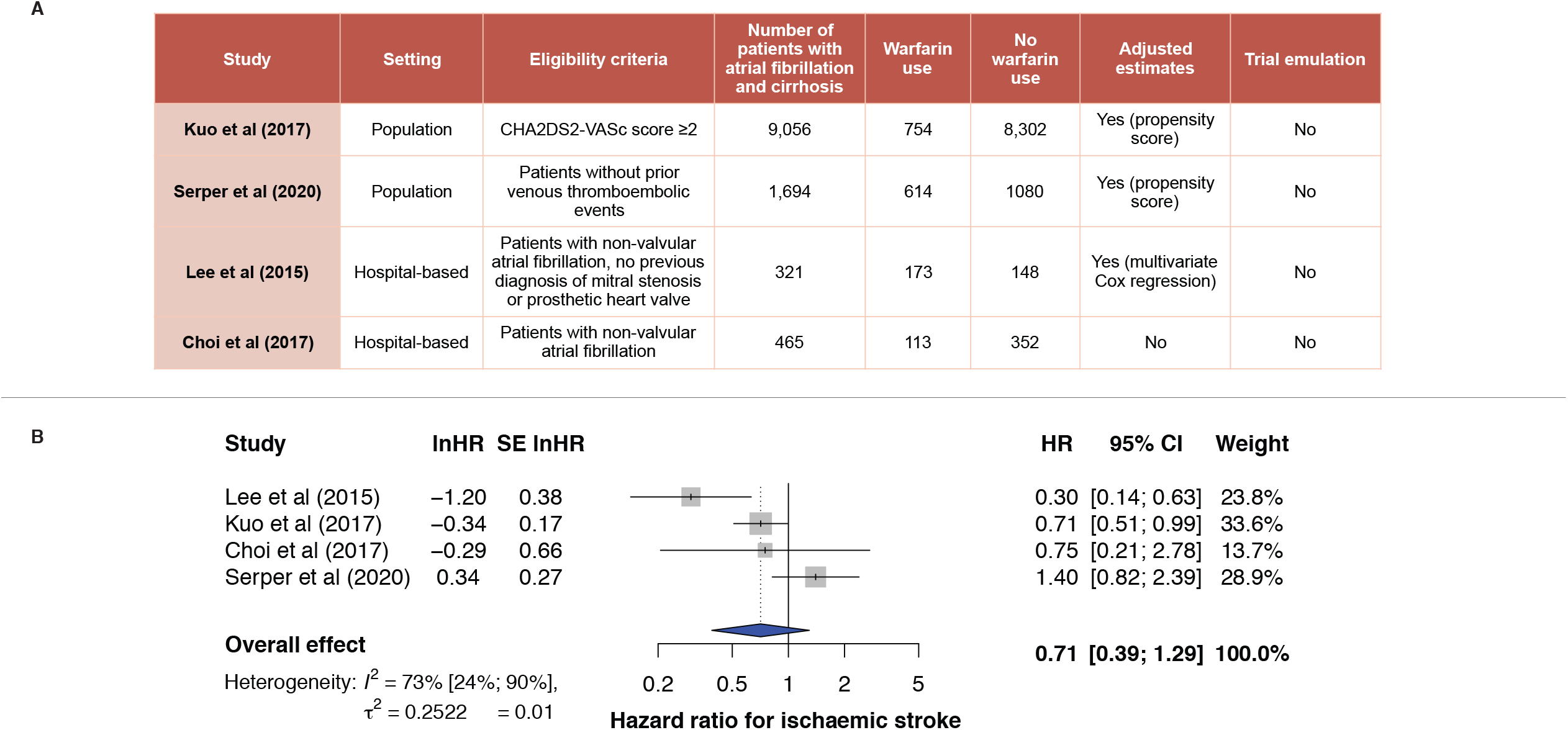
Informatics Consult Report: New synthesis of prior observational evidence. Meta-analysis of the association between warfarin use and the risk of ischaemic stroke in observational studies including approaches for automation. (A) Characteristics of observational studies included in the meta-analysis. (B) Forest plot depicting the hazard ratios calculated with the DerSimonian and Laird random-effects models. HR = hazard ratio; CI = confidence interval; SE = standard error.

### Target trial emulation using population-based EHRs

Per the target trial protocol, a cohort encompassing 1,022 individuals fulfilling all eligibility criteria was created (initiators = 443; non-initiators = 579). We performed PSM on 22 baseline covariates and generated a matched cohort involving 235 initiators and 526 non-initiators (Figure 5A), baseline patient characteristics before and after PSM are shown. We estimated an intention-to-treat HR for all-cause mortality of 0.61 (CI = 0.49 – 0.76; p < 0.0001) comparing warfarin initiators with non- initiators (Figure 5B). Warfarin used was associated with lower risk of ischaemic stroke: HR = 0.27 (CI = 0.08 - 0.91, p = 0.034) (Figure 5B). The 1,022 eligible participants for the target trial were categorised into the five subgroups (aged ≤ 65, aged > 65, men, women and patients with normal INR), followed by PSM. Intention-to-treat HRs for all-cause mortality comparing warfarin initiators versus non-initiators were as follow: aged ≤ 65: HR = 0.62 (0.45 - 0.86, p = 0.0041); aged > 65: HR = 0.61 (0.46 - 0.83, p = 0.0015); men: HR = 0.64 (0.49 - 0.84, p = 0.0014); women: HR = 0.53 (0.37 - 0.77, p = 0.00087) and normal INR of < 1.7: HR = 0.62 (0.50 - 0.78, p < 0.0001), where warfarin therapy is associated with lower mortality risk.

**Figure 5.**
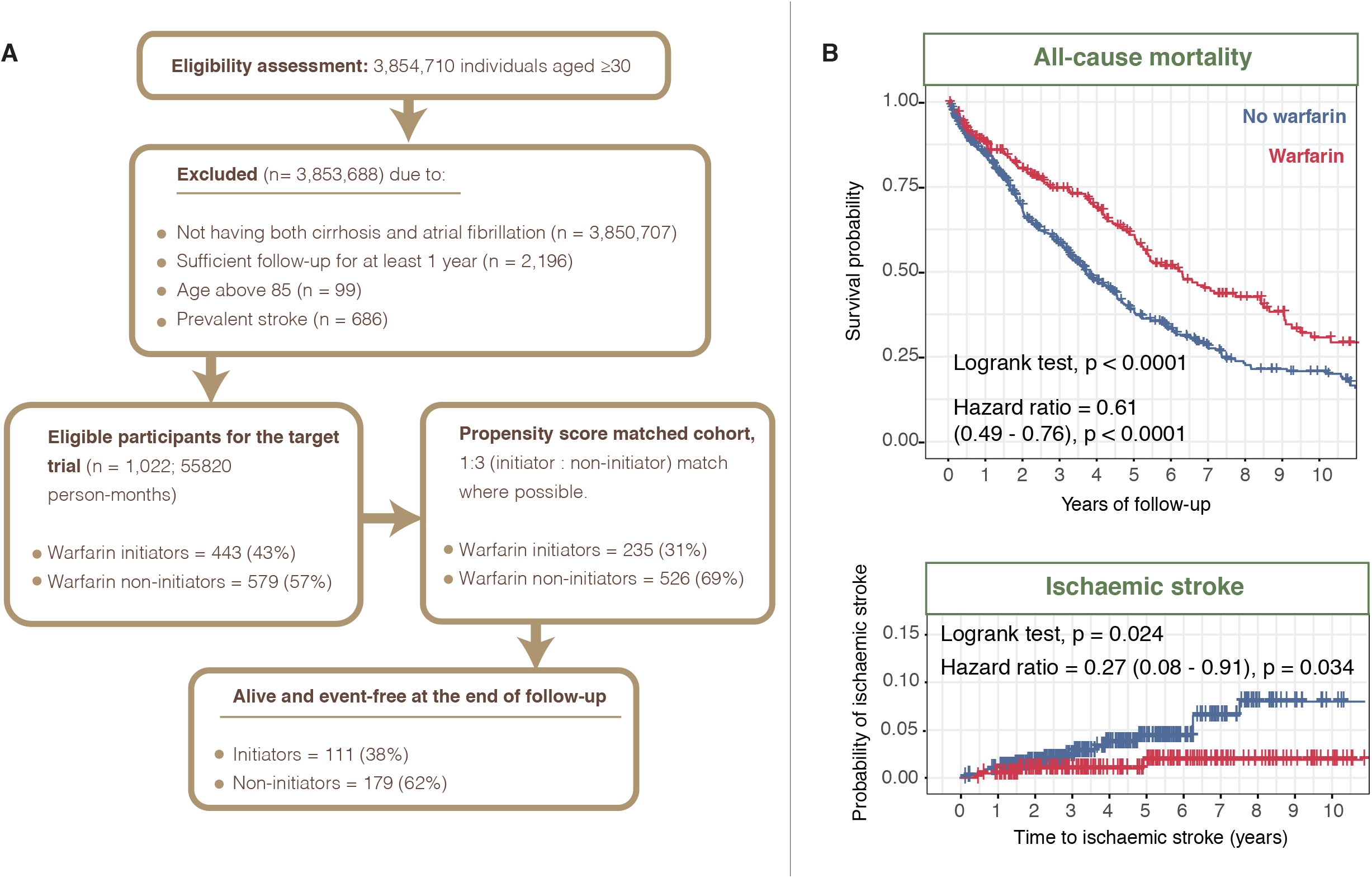
Informatics Consult report: New observational evidence through target trial emulation (intention-to-treat analysis) where eligibility and treatment assignment were aligned with time zero of follow-up, as is done in randomised controlled trials. (A) CONSORT diagram showing the selection of eligible individuals for the target trial emulation of anticoagulation therapy in patients with atrial fibrillation and cirrhosis. (B) Kaplan-Meier plots of the propensity-matched cohort for all-cause mortality and ischaemic stroke. Flow diagram depicts analysis design. P values from logrank tests were indicated. Hazard ratios from Cox proportional hazards regression analyses were indicated. Numbers in parentheses indicate the 95% confidence intervals.

### Genetic evidence

We found no GWAS summary data for vitamin K1 in patients with atrial fibrillation and cirrhosis, but we did find genetic evidence to indicate that warfarin use is associated with reduced stroke risk. For two-sample MR, we used four methods (inverse-variance weighted (IVW), MR-Egger, simple median and weighted median). The IVW analyses, which assumes no pleiotropy, revealed that higher genetically predicted levels of vitamin K1 were associated with a higher risk of any stroke with an odds ratio (OR) of 1.06 (95% CI: 1.00 - 1.11) per Ln-nmol/L increase in vitamin K1 (Figure 6). However, these results were not replicated using methods (simple median, weighted median and MR-Egger) which allow for genetic pleiotropy. When considering stroke subtypes, we observed that higher genetically predicted levels of vitamin K1 were associated with a higher risk of large artery atherosclerotic stroke for 3 out of 4 MR methods: simple median (OR = 1.25 [1.03 - 1.51]); weighted median (OR = 1.25 [1.03 - 1.52]) and IVW (OR = 1.29 [1.11 - 1.50]) (Figure 6).

**Figure 6.**
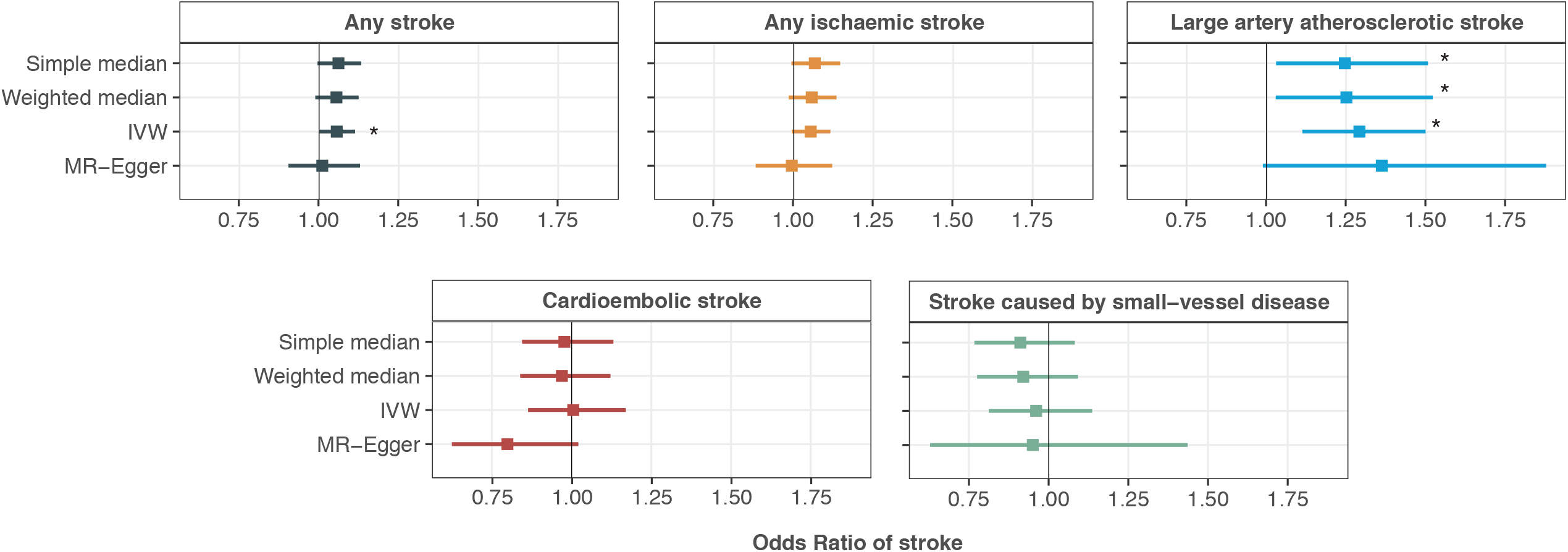
Informatics Consult report: Detailed genetic evidence.Two-sample Mendelian randomisation on circulating vitamin K1 levels and risk of stroke. * indicates significant results.

### Timely report generation

Generating the evidence and report took one month (two analysts from scratch, without having previous protocols to follow). EHR phenotypes were, however, already created, validated and implemented which sped up the process significantly (https://portal.caliberresearch.org/)[17]. Table 1 shows additional opportunities for pipelining each of the four streams of evidence identification and generation to return the Consult report. Computable EHR phenotypes and computable clinical trial protocols can be used to automate the process of trial identification and trial recruitment[25,26]. For meta-analyses of observational studies, approaches for semi-automated systematic reviews[27,28], batch extraction of data from articles[29,30] and mapping of SNOMED-CT terms to MeSH descriptors in PubMed[31] can be used in the pipelining process. Automating these steps and performing data extractions from articles in batch mode will enable the return of results within 24 hours. The DExtER tool can be used for automatic extraction of EHR databases and automated cohort creation for the target trial emulation process[32]. Recent addition of an analytical module to DExtER enables cohort creation, statistical analyses and results visualisation within short time scales (1-4 hours) for matched cohort studies. Ongoing work will make DExtER return results real- time. Two-sample MR to generate genetic evidence can be automated using the MR-Base platform[33], which returns results within minutes. By using such approaches to automation, we estimate all four streams of new evidence could be generated in 48 hours by one health informatician from the point at which the clinician makes the Consult request.

**Table 1:**
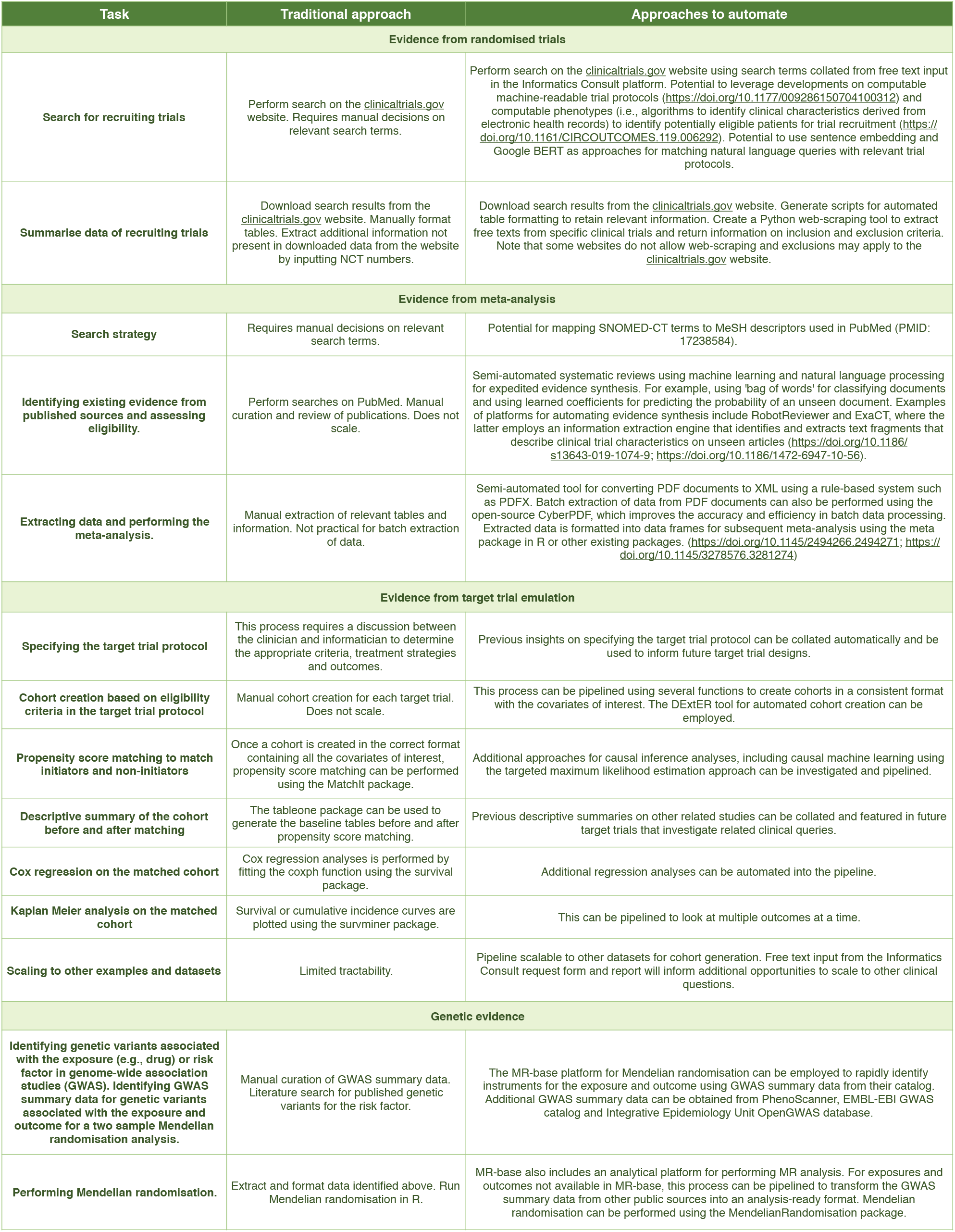
Traditional versus automated approaches for evidence synthesis.

### Clinician survey on the acceptability and feasibility of the Informatics Consult

We surveyed an independent sample of 34 clinicians from eight specialties with results shown in (Figure S2). Results indicated that 79% of clinicians thought that they should have access to the Informatics Consult as a service within their healthcare systems (21% responded ‘maybe). Clinicians found each section of the report useful (or ‘maybe useful’) as follows: prognosis 85% (12%), a summary of evidence of efficacy and safety 79% (21%) and disease prevalence 68% (18%). Only 18% of clinicians thought that they needed to have access to the details of the evidence in the clinic with one clinician stating: “*Multi-disciplinary team meetings might benefit from Informatics Consults, this is where difficult cases are discussed and there is time to review newly generated evidence*”. When asked whether clinicians found detailed reports on the four sources of evidence useful (or ‘maybe useful’), responses were as follow: randomised trials: 82% (15%); meta-analysis of prior observational evidence: 76% (18%); target trial emulation: 62% (18%) and genetic evidence: 26% (24%). Clinicians would discuss the Informatics Consult report with their patients 74% (26% answered maybe). Clinicians offered 15 further clinical questions where the Informatics Consult might be of value and made additional comments including: “*Ultimately, we practise defensive medicine - would my decision stand up in court based on available data – the Informatics Consult should help with that*.” and “*Regulators and guideline developers will require replication and quality assurance of evidence generated in clinical timescales*”.

## Discussion

We demonstrate that the Informatics Consult offers a novel paradigm to generate new clinical evidence and return results within clinical timescales. We found that in patients with atrial fibrillation and cirrhosis, initiation of warfarin was common, may be associated with lower all-cause mortality and may be effective in lowering stroke risk. Given the ubiquity of clinical uncertainty where there is little or no evidence, and that current modes of generating new evidence may never be initiated or, if initiated, take years to report, it is imperative to accelerate learning from extant data.

### Informatics Consult versus traditional approaches for evidence generation and delivery

The Informatics Consult puts the treating clinician and the patient at the centre of evidence generation. Indeed, in seeking to address a range of questions from the clinician and patient, the Informatics Consult enables simultaneous delivery of evidence from different sources, rather than employing a one-study-one-design-at-a-time approach. The Consult is embedded within EHR system and evidence is generated within clinical timescales—making it a form of an electronic consult, which is increasingly being adopted to seek specialist input[34,35]. Additional information on how the Informatics Consult differs from traditional approaches to evidence generation and use is summarised in Table S1.

### Concordance across sources of evidence identified from the Consult

A primary motivation for requesting an Informatics Consult is to understand how a particular treatment influences an outcome to help guide decision making. We show a degree of concordance across four sources of evidence. RCTs demonstrate the effectiveness of oral anticoagulation in stroke prevention in patients with atrial fibrillation: but as we demonstrate reported and currently recruiting RCTs exclude patients with cirrhosis (i.e., patients having ‘contraindications’ for anticoagulants). This suggests that the prospect of ever mounting, or successfully recruiting to, an RCT in patients with AF and cirrhosis is low. Meta-analysis of observational studies and target trial emulation suggest evidence on the potential benefits of warfarin for stroke reduction, suggesting the significant impact on strokes and deaths averted if these patients are treated with anticoagulant therapy. Although we did not find any relevant GWAS summary data, evidence suggests that lower levels of vitamin K1 (target of warfarin) are associated with lower stroke risk which is corroborated by another study[36].

### Returning results on prevalence, prognosis and treatment variation

Interestingly, the clinicians involved in this study recommended that information beyond efficacy and safety should be included as options for the clinician to request in the Informatics Consult. By providing information on the prevalence of cirrhosis and atrial fibrillation, we demonstrate that this pair of conditions is not highly prevalent (although not considered rare). Knowing that the health system has data on diagnosis, treatment and outcomes in an estimated 35,000 individuals (scaled up to the population of England) with both conditions, highlights the importance of several areas. First, the importance of accessing and learning from nationwide data at scale; any one clinician may have clinical experience of only a handful of such cases, and it is not feasible or scalable to create registries. Second, the clinician likely has never had access before to population-based, contemporary prognostic information on such patients (1-year mortality: 15%), nor the knowledge that in practice 43% of patients were started on warfarin. We anticipate that providing clinicians and patients with this information may stimulate further questions on generating new Consults on a wider range of prognostic outcomes, and predictors of prognosis.

### Need and demand for Informatics Consult

Nearly all clinicians are faced with treatment decisions where limited evidence exists where insights might be gained from analysis of large-scale data on ‘patients like me’. But currently, few if any clinicians can request such insights. A clinician from the survey mentioned that evidence from the Informatics Consult is important to help their decision in practicing defensive medicine given that the majority of clinical practice recommendations in professional society guidelines are not supported by RCT evidence. There is limited information on the system-wide frequency of treatment ‘clashes’ where indication and contraindications coexist in the same patient. This is especially relevant for patients with multimorbidity where evidence from RCTs are limited[6], which may result in individuals being subjected to low-quality recommendations. For example, certain targeted cancer therapy such as angiogenesis inhibitors may cause an increase the prevalence of hypertension during treatment, which highlights the importance of considering the impact of cancer therapy on adverse side effects and cardiotoxicity[37]. Our preliminary analyses demonstrate that 26 in 10,000 women have breast cancer and hypertension; these individuals might benefit from the Informatics Consult.

### Developing the Informatics Consult as a service

The majority of clinicians in our survey thought that they should have access to the Informatics Consult as a service as their healthcare systems seek to learn from existing data[38]. We have demonstrated the feasibility with this one example. Scaling to other clinical examples, and a service, requires five inter-related challenges to be addressed in clinical standards setting, implementation and evaluation, access to data, informatics, and knowledge management. First, bodies that define standards of care – including clinical practice guideline developers and regulatory and technology assessment bodies – are already considering real-world evidence generated in conventional timescales[7,39]. In the context of a more rapid generation of evidence, the Informatics Consult raises new questions about replication, open peer review and quality assurance pipelines. Second, development, implementation and evaluation of the impact of the Informatics Consult on clinical decision making in practice are required. Feedback from our survey suggests that Multidisciplinary Team (MDT) meetings, which concentrate on ‘difficult’ clinical cases may present an opportunity[40]. Third, an Informatics Consult service requires approved, immediate access to large scale, clinically detailed updated data, most likely in a trusted research environment. The coronavirus pandemic offers a new precedent for such information governance approvals, which can not foresee the nature of the next clinician’s question. Fourth, biomedical knowledge (in EHRs, trial protocols, clinical guidelines) needs to become more computable and interoperable. Design and analysis methods need to be improved, standardized and widely distributed. For example, the Observational Health Data Science and Informatics community offers open-source software implementing many routine analyses methods[41]. It took one month to generate simultaneously the findings on four streams of evidence. Without the Informatics Consult, this evidence may never be generated at all; or may be published at different times over a period of years. A feasible goal, using automation, would be to generate the report in 48 hours. Fifth, there needs to be an open process of making the knowledge available in an Informatics Consult library. As the library builds, Clinicians might make a request for which a previous consult already provides an answer. The Library might also serve as a platform for connecting ‘patients like me’, registering the frequency of therapeutic dilemmas and potential treatment uncertainties, identifying the need for new RCTs, and informing their design and facilitate targeted recruitment into trials.

### Strengths of the study

To the best of our knowledge, this is the first demonstration of Informatics Consult for a treatment decision, triangulation of evidence from meta-analyses of observational studies, target trial emulation using EHR data and MR. The breadth, depth and longevity (long follow-up period) of the EHR data, which links primary care, secondary care and the death registry is an advantage. Another significant strength is engagement with an independent sample of clinicians from multiple specialties to gauge the feasibility and acceptability of the Informatics Consult.

### Limitations of the study

Our study has important limitations. First, although we have prototyped EHR request and report forms based on feedback from clinicians, these have not yet been implemented in live clinical systems. Second, as initial proof of concept, we have not assessed bleeding outcomes and newer non-vitamin K oral anticoagulants due to current data access limitations. Third, it remains difficult to assess whether there unmeasured confounding despite employing causal inference methods. It is also possible that some prescription data is missing, which means that patients are incorrectly assigned to the control arms. Future analyses on a more recent and larger dataset involving both CPRD Gold and Aurum[42] would be beneficial. Fourth, a limitation of the MR analyses is that access to summarised GWAS data for stroke outcomes in patients with cirrhosis is limited. The advantages of using publicly available GWAS summarised data are speed and transparency, both of which are essential to the Informatics Consult. Although individual-level data would allow more flexibility to conduct analyses in specific patient subgroups and to select which variables to generate the summarised data for, such analyses could not be returned within a clinical timescale that is not scalable and cannot be fully automated. Fifth, although we have identified potential ways to automate the analytic process, these have not been implemented here.

## Conclusion

We proposed an Informatics Consult framework to summarize evidence from four sources and have developed a report prototype for answering a treatment question to enable new ways of data- informed decision making in clinical timescales. The Informatics Consult may stimulate a conversation among public, professionals and policymakers about more rapidly realising the benefits of health system learning from ‘patients like me’.

## Data Availability

All data is shown in the manuscript.

## Acknowledgements

We thank clinical colleagues who have contributed to the pilot survey and who have consented for their names to be acknowledged: Christopher Tomlinson, Upkar Gill, Paul Harrow, Harry Martin, Omer Ahmad, Louise China, Vanessa Taylor, Sai Ambati, Amy Prideaux, Alex May, Ruth Gilbert, Hamish Miller, Bilal Mateen, Rhys Davies, Bu’Hussain Hayee, David Cronin, Katharine Pollock and Philip Oppong.

## Authors’ contributions

Research question: AGL, GRF, HH Funding: AGL, HH

Study design and analysis plan: AGL, WHC, CP, HH

Preparation of data, including electronic health record phenotyping in the CALIBER portal: AGL, WHC, CP, SD

Statistical analysis: AGL, WHC, MK

Drafting initial versions of the manuscript: AGL, HH

Critical review of early and final versions of the manuscript: All authors

---

**Figure S2.**
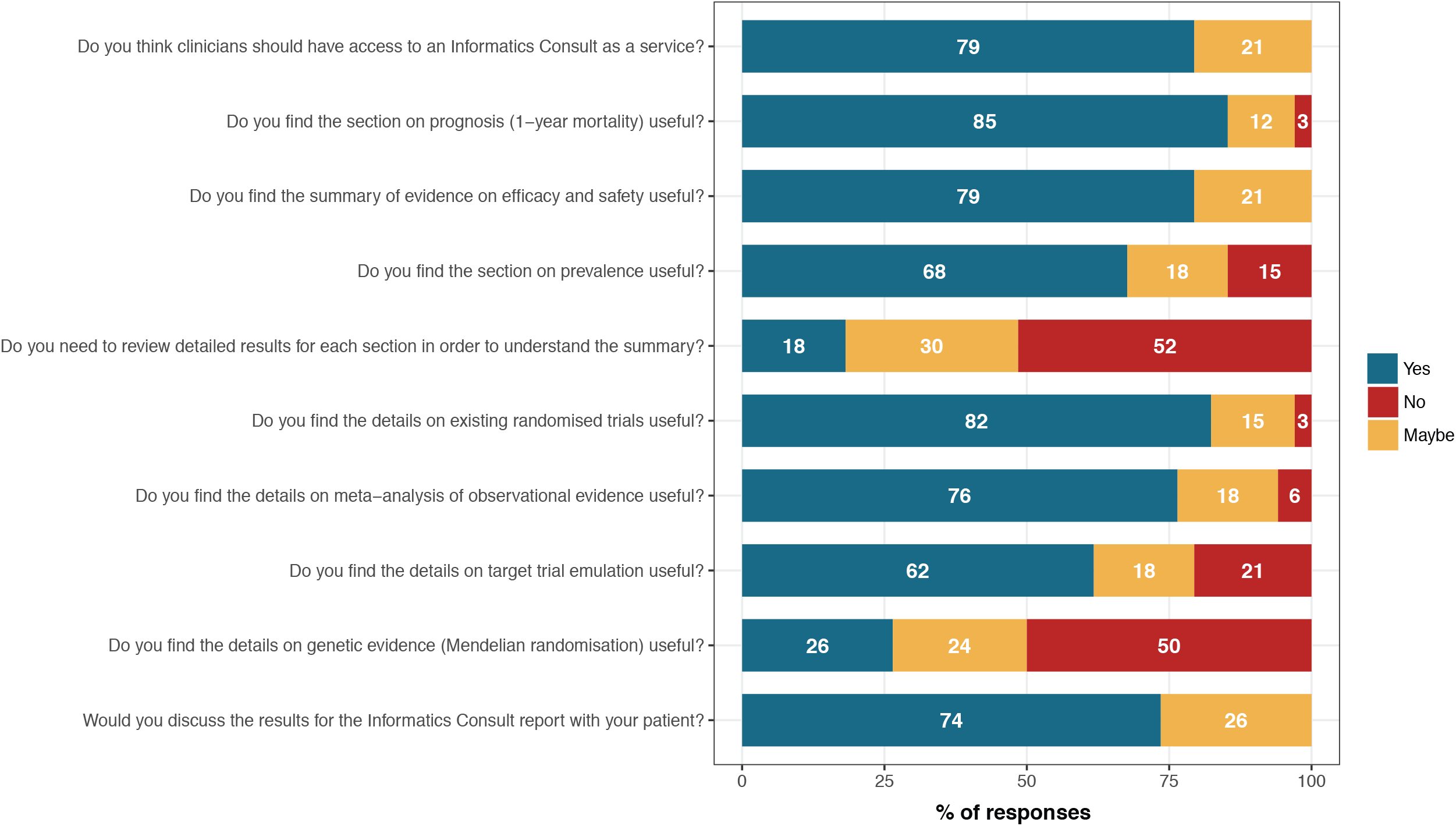
Summary of survey responses.

**Table S1:** Comparison of the Informatics Consult with traditional approaches to evidence generation.

| Areas | Traditional methods of evidence generation | Informatics Consult |
| --- | --- | --- |
| Who initiates the question? | Top down: typically driven by a small number of researchers | Bottom up: democratic, any clinician, any uncertainty and potentially patient driven |
| Simultaneous delivery of all relevant evidence | Not often possible, one study and one design at a time | Yes |
| Timescale for generating evidence | Months or years from question to answer | Hours, within clinical timescale |
| Embedded in health systems (the question or the answer or the means of providing answer) | No | Yes |
| Clinical trial prioritisation, informing trial design and recruitment | Top down: driven by specific groups of individuals | Bottom up: opportunities for embedding within clinical practice |
| Involvement of individuals | Researcher centric | Patient and clinician centric |
| Strength of evidence and levels of evidence | Levels of evidence classified in all major clinical guidelines; however, these are all based on peer reviewed published evidence | Same levels of evidence apply; however, the framework aims to return evidence within clinical timescales to mitigate hurdles in the journal peer-review process. Potential for collating information from past Consults in an open-access repository. |
| Clinical guideline recommendations based on evidence | Yes, information from randomised trials and peer-reviewed publications are included in clinical guidelines | Raises important questions on what basis might clinicians follow the Consult before it is peer reviewed and published |
| Regulatory approvals on the labels of drugs and medical devices | Food and Drug Administration (FDA), European Medicines Agency and Medicines and Healthcare products Regulatory Agency established processes for trials and for the Real-world Evidence framework | Largely untested |

